# A Framework for Assessing the Impact of Accelerated Approval

**DOI:** 10.1101/2022.02.14.22270951

**Authors:** A. Lawrence Gould, Robert K. Campbell, John W. Loewy, Robert A. Beckman, Jyotirmoy Dey, Anja Schiel, Carl-Fredrik Burman, Joey Zhou, Zoran Antonijevic, Eva R. Miller, Rui Tang

**Affiliations:** Methodology Research, BARDS, Merck & Co., Inc., Kenilworth NJ, USA; Molecular Pharmacology, Physiology and Biotechnology, Brown University, Providence RI; DataForethought, Winchester, MA, USA; Lombardi Comprehensive Cancer Center and Innovation Center for Biomedical Informatics, Georgetown University Medical Center, Washington, DC, USA; Data and Statistical Sciences, AbbVie, North Chicago, IL, USA; Department for Pharmacoeconomics, Norwegian Medicines Agency, Oslo, Norway; Data Science & AI, AstraZeneca R&D, Gothenburg, Sweden; Xcovery Pharmaceuticals, Palm Beach Gardens, FL, USA; Abond CRO, Allendale, MI, USA; Independent Biostatistical Consultant, Middletown Twp, PA, USA; Methodology and Data Visualization, Biostatistics Department, Servier Pharmaceuticals US

**Keywords:** Decision Analysis, Regulatory Science, Benefit/Risk

## Abstract

The FDA’s Accelerated Approval program (AA) is a regulatory program to expedite availability of products to treat serious or life-threatening illnesses that lack effective treatment alternatives. Ideally, all of the many stakeholders such as patients, physicians, regulators, and health technology assessment [HTA] agencies that are affected by AA should benefit from it. In practice, however, there is intense debate over whether evidence supporting AA is sufficient to meet the needs of the stakeholders who collectively bring an approved product into routine clinical care. As AAs have become more common, it becomes essential to be able to determine their impact objectively and reproducibly in a way that provides for consistent evaluation of therapeutic decision alternatives. We describe the basic features of an approach for evaluating AA impact that accommodates stakeholder-specific views about potential benefits, risks, and costs. The approach is based on a formal decision-analytic framework combining predictive distributions for therapeutic outcomes (efficacy and safety) based on statistical models that incorporate findings from AA trials with stakeholder assessments of various actions that might be taken. The framework described here provides a starting point for communicating the value of a treatment granted AA in the context of what is important to various stakeholders.

## Introduction

The FDA’s Accelerated Approval program (AA) is one of four FDA programs to expedite availability of products that treat serious or life-threatening illnesses where effective treatments do not exist or where currently available treatments are inadequately effective or unacceptably toxic (Appendix 1). A product is a candidate for AA if it meaningfully affects a surrogate endpoint that is *reasonably likely* to predict clinical benefit or a clinical endpoint that can be measured earlier than irreversible morbidity or mortality (IMM) and is reasonably likely to predict a subsequent effect on IMM.[1]

From 1992 through mid-2021, the FDA granted 269 AAs for 162 different treatments (drugs or drug combinations). [2] Most were for infectious disease (24/162) and cancer (113/162). The creation of the AA pathway was influenced by the HIV epidemic and patient advocacy. Thirty HIV treatments received accelerated approved using surrogate endpoints and all fulfilled their post-approval study requirements.[3] The surrogate endpoints for efficacy against HIV proved to be highly predictive of clinical outcome, and were eventually adopted for traditional approval. These drugs and post-approval innovations in their use enable HIV-infected persons to have life expectancies close to the life expectancies of uninfected persons.[4] There have been more than 110 AAs for malignant hematology and oncology indications.[5] The emergence of targeted cancer drugs and immune checkpoint inhibitors was accompanied by increased use of AA in oncology, with 70 approvals from 2014-2018.[5] Six oncology AAs granted before 2009 were withdrawn, with three of the drugs resubmitted and approved based on improved evidence supporting their use. [6] A seventh withdrawal was initiated in 2019.[7] Almost all of the cancer AAs prior to 2016 have been converted to full approvals, while most from 2016-2018 remain to be converted.

All stakeholders (patients, physicians, sponsors of innovative treatments, reimbursement agencies, and regulators) ideally should benefit from AA. However, concern has been expressed about the value of accelerated approvals.[8] Some of the main points of conflict are over what has been measured in studies supporting accelerated approval, who has been assessed in these studies, and how long it will take for results to become available for conversion to traditional approval. AA trials provide less information than regulatory approval usually requires, and the short-term benefit/risk balance based on these trials may not reflect the true benefit/risk balance for the product. Decision-makers must rely on evidence that is less complete than what current practices provide to create cost-effectiveness assessments of new treatments.[9] Although the regulations concerning AA require sponsors to conduct clinical trials to establish efficacy based on clinical benefit, adherence in this regard has not been universal.[10] Issues affecting the completion of these requirements include equipoise concerns when a drug has been shown superior to standard care in the AA trials [11] and reluctance of patients to participate in a randomized confirmatory trial of a marketed drug available without the risks or inconvenience of research.

A common definition of the intuitively appealing term ‘impact’ that is used in discussions of the benefits and risks of AA can be elusive. There are many possibilities, depending on the needs and values of various stakeholders. For this discussion, impact refers to the value of suitable metrics reflecting subsequent actions that a stakeholder might take as a result of an evaluation of the information supporting AA. For example(s), a regulatory agency may decide to approve or not approve a drug via AA depending on the distribution of possible predicted metric values such as estimated probability of survival for more than 5 years regardless of promising early findings. An HTA could decide to put a drug approved via AA on its formulary for reimbursement (or not) depending on the distribution of possible predicted metric values such as whether the anticipated cost per quality-adjusted life year was within an acceptable range. A sponsor might decide whether or not to go forward with seeking AA given the metric value findings from trials that might be submitted in support seeking AA. Physicians and patients might decide to initiate treatment (or not) depending on possible predicted outcomes. “Values” in this context depends on what is important (and how important) to each stakeholder. Stakeholder values would be elicited in any implementation. Different stakeholders will have different attributes that are important to them, and even may differ in how importantly they regard attributes that are shared with other stakeholders.

An evaluation of the impact of AA is essential for assessing its value as it applies to the stakeholders that it affects. Values for stakeholders can be unique or shared, as illustrated conceptually in Figure 1. A common process for evaluating the impact of AA can be helpful in integrating the assessments reflecting the values for the various stakeholders. These values can provide useful guidance for decisions that any stakeholder might consider.

**Figure 1.**
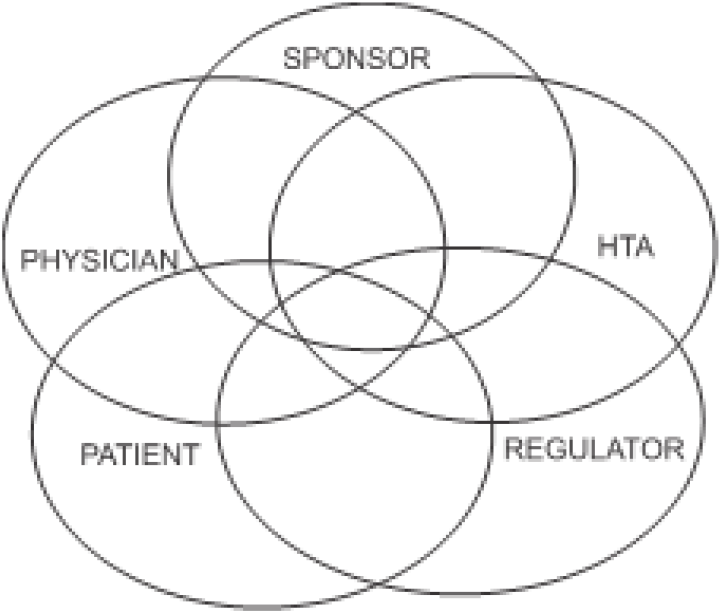
Unique and shared values of key AA program stakeholders.

A recent white paper from the Institute for Clinical and Economic Review [12] proposes strategies for strengthening the performance of the AA process by addressing a number of concerns that are relevant for stakeholder assessment once AA has been granted. These concerns include inconsistencies in the level of uncertainty deemed to qualify surrogate endpoints as reasonably likely to predict a clinically meaningful treatment effect; a lack of clarity over what magnitude of change in such outpoints justifies accelerated approval; and the high prices commanded by these products despite their relative lack of evidence.

We propose a common framework and, therefore, a common language, for addressing the challenges of decision making faced by various stakeholders inherent with the Accelerated Approval process that differs from decision science based methods that have been described previously. With this proposal, each stakeholder uses information from the AA trials to decide on a course of action that reflects stakeholder-specific values by an objective, transparent, and reproducible process that differs among stakeholders only with respect to the stakeholder-specific actions and values. This approach provides a way for stakeholders to understand the bases for each other’s decisions and may be useful for informing their own decisions. The specificity of the values for different stakeholders means that the stakeholders could reach different action decisions, possibly at different points in time, so that there may be no immediate determination of ‘consensus’. Consequently, the common framework approach described here differs from the consensus-based decision model for assessing health systems described by Xu *et al* [13] in which different statistical decision methods employed by stakeholders with a common objective are integrated to reach a consensus. It also differs from previous applications of decision science methods that focus on sponsor decisions about confirmatory or post-POC study designs, and how these design options could affect outcomes, utility, and future (post-approval) decisions by stakeholders.[14-18]

The context differs as well from previous work in that not all stakeholders implement their decision processes contemporaneously. Accelerated Approval trials provide much drug-related (and competitive) evidence that can affect some stakeholder decisions. This evidence includes the study results, label content, FDA review documents (sometimes), and the current treatment landscape. Most importantly, since the drug has been approved to market (at least in the US), the decisions at issue are pushed from the sponsor and regulatory agencies to stakeholders such as patients, physicians, and health technology agencies that determine implementation in clinical care.

## Method

### Defining impact

This article outlines a process for defining ‘impact’ that is grounded in well-established decision theory. Although the process is objective and reproducible, its components may be objective or subjective, to reflect clinical reality, and the implementation details can be adapted to the needs of different stakeholders. For clarity of exposition, the description that follows focuses on the impact of AA on patients who receive the drug through access enabled by accelerated approval. Similar developments can be carried out for the other stakeholders. Decision theoretic concepts reflecting the values of different stakeholders have been applied to the design and analysis of clinical trials when only a subset of the patient population may be likely to benefit materially from the test drug. [14, 17-19]

There are at least two key assumptions and two parts to the process. The first key assumption is that the expected <u>clinical</u> benefit of a treatment for a patient is a function of the short-term outcomes observed in the studies supporting AA, observable patient attributes, and, possibly, unobserved confounding factors. The assumption implies that the short-term benefits on biomarkers or surrogate endpoints demonstrated by these studies would predict reliably (in a probabilistic sense) the actual clinical benefits and risks for a patient undergoing long-term treatment. This could be questionable for various reasons, especially when there is a substantial lapse of time between the accelerated and the clinical endpoints. The anticipated clinical benefit generally will depend on the nature of the disease itself and the expectations of the treatment, whether short-term or long-term, including the usually disease-specific therapeutic strategy.

The second key assumption, common to all clinical development programs, is that patients are partially or wholly exchangeable [20] so that the same model applies at least for important subgroups of the patients (e.g., men, women), certainly for those in the AA trials and ideally for all patients who might be treated.

The first part of the process entails estimation of the parameters of models relating clinical responses/outcomes resulting from the innovative treatment to the patient attributes and short term findings from AA studies, including assessments of the variability of the parameter estimates and the uncertainty about predicted clinical outcomes. Ideally, if (a) the general functional (mathematical) relationship between the attributes of a patient and the clinical response is known, (b) the exact (error-free) values of the parameters that characterize the functional relationship as it applies to a particular patient are known, and (c) there are no confounding factors then, at least in principle, it would be possible to determine the expected clinical outcome for any patient. In reality, the model and parameter values are not known exactly, although reasonable approximations may be achievable based on available data. However, even if one could determine the expected clinical outcome exactly for any patient, intrinsic variability among patients would lead to different actual responses of patients with the same expected response, so that parameter values still would need to be estimated.

The second part of the process is the use of the parameter estimates to predict the efficacy and toxicity outcome (clinical response) for a patient who is a candidate for treatment with a product that received AA. This prediction is subject to uncertainty due to intrinsic patient variability, the uncertainty associated with the estimates of the parameters used to determine the expected response, and the influence of possible confounders.

Evaluations of the impact of AA do not take place in an innovative vacuum. The overall benefit of a therapy in a given population will depend on how long it remains in use before it is replaced by superior therapeutic alternatives.[19, 21] The therapeutic alternatives at the time of accelerated approval will influence the initial impact of the AA, while subsequent approvals of other therapies for the same indication and patient population will impact the overall time course of this impact

### Impact on patients

The potential impact for a patient treated with a product receiving AA could be positive if the product conveys clinical benefit relative to the best available treatment option for the patient, or negative if it prevents the patient from receiving a better treatment or makes the patient’s condition worse because of toxicity. Many other factors influence the impact on a patient, particularly those affecting quality of life such as discomfort, management of dosing, lost work hours, burdensome medical costs, and paperwork management. These can in principle be incorporated in the assessment of impact, although specifically how would depend on the particular circumstances. Not treating the patient with a truly effective product could have a negative impact because the patient would fail to achieve the product’s benefit while avoiding potential toxicities of alternative treatments. Ideally, AA provides information for estimating the probabilities corresponding to possible therapeutic outcomes for a patient that can be refined as more information accumulates. Eichler *et al* [22] describe how this might be carried out.

An unintended negative impact of granting AA could be the failure of an ongoing trial of a potentially effective agent because of patient reluctance to be randomized so that future patients may not receive the drug because a reimbursement body (e.g. IQWiG [Institut für Qualität und Wirtschaftlichkeit im Gesundheitswesen] in Germany) could conclude that insufficient evidence of benefit had been shown. Consequently, it could be more beneficial to refuse an AA application if full approval could be granted shortly afterwards when longer term data are available.

A key objective of this article is the proposal to assess the impact of AA in an objective, transparent, quantitative, and reproducible way by means of well-known statistical decision principles. Appendix 2 briefly outlines the statistical decision process used here; detailed descriptions of the theory and principles are available in the literature. [23-26] Realistic models could incorporate more detailed specification of what might be meant by ‘outcome’, effects of possible selection bias based on promising early results, the choice of endpoints, consideration of the influence of possible differences between the populations in the AA trials, in the confirmatory trials. and in general clinical practice. At best, caution is required because the findings from one trial may not be replicated in a second trial or in a general clinical population. [27-29]

### Considerations for assessing impact

The efficacy and toxicity outcomes of a therapy are important considerations for assessing the impact of AA, but not the only ones; different stakeholders may focus on different issues such as benefit/risk or cost considerations. What follows focuses for simplicity of exposition on efficacy and toxicity issues. Suppose for example, that the short-term efficacy information from the AA trials defines a predictive distribution of a relevant clinical response (e.g., progression-free survival past 6 months), and that a similar distribution of clinical response can be determined for a previous therapy. Figure 2 displays some possibilities for the distributions of the clinical response for the new and previous therapies, including the possibility that the distribution of clinical responses for the new therapy may be reflect greater variability than for the previous therapy because of less information about the clinical responses for the new therapy.

**Figure 2.**
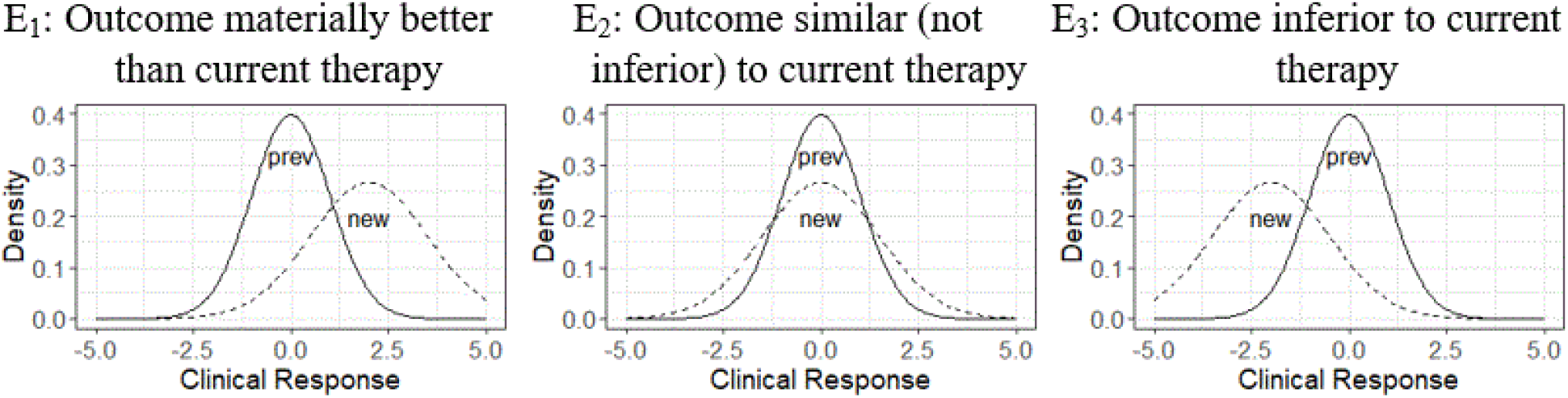
Orderings of clinical response for a new and a previous therapy.

The orderings of the clinical response distributions can be expressed by various metrics. Basing the orderings of efficacy (and toxicity) on the predictive distributions of the outcomes reflects uncertainty both about the actual AA outcomes and the basis for inferring the likely clinical outcomes based on the AA results.

Suppose that the efficacy information provided in support of AA uses patient and treatment attributes to map the predicted efficacy outcomes (E = [E_1_, E_2_, E_3_]) to the categories/ conclusions illustrated in Figure 2. These categories refer to the desired efficacy outcomes, not to the metrics used to support AA. The mapping depends on the indication, the disease, clinical judgment, and the expectations of therapy.

The predicted toxicity findings similarly can be assigned to various categories as in Table 1. ‘Better’, ‘outcome’, and ‘toxicity’ can mean different things to different people/stakeholders. We focus here on their meaning <u>from the patient’s point of view</u>. Suppose that probabilities associated with the various efficacy and toxicity categories can be related to predictive information about a patient who could be, but has not yet been, treated with the product, and the condition to be treated. The prediction depends on the attributes of the candidate patient and the parameters of the model that relates these attributes (including treatment) to the anticipated clinical outcome. The predicted outcome is subject to uncertainty about the true values of the parameters, including the accuracy of the prediction of clinical outcome based on surrogate measures, and intrinsic variation among patients with identical attributes. This simple approach can be extended in various ways including incorporating time into the evaluation of the effect of AA.

**Table 1.** Predicted toxicity outcome.

|  |  |  |
| --- | --- | --- |
| T = | T<sub>1</sub> : | Less toxic than current therapy |
|  | T<sub>2</sub> : | About as toxic as current therapy |
|  | T<sub>3</sub> : | More toxic than current therapy, but manageable |
|  | T<sub>4</sub> : | Intolerable toxicity |

These outcome categories can be combined into clinically meaningful categories describing predicted therapeutic outcomes for patients that combine efficacy and toxicity. These categories ordinarily will be defined in numerical terms such as expected duration of progression-free survival so that, for instance, a better outcome could be a lower confidence bound on the hazard ratio for the new treatment relative to the standard that exceeded 1.5 (i.e., 50% improvement in survival). What follows assumes that there are appropriate definitions of categories of efficacy and toxicity. Let y_P_ denote the predictive observations made on a candidate for treatment with the product in question. The surrogate endpoint and biomarker values obtained in the AA trials may be useful for determining predictive distributions of therapeutic outcomes and, consequently, estimates of the probabilities associated with each of the Efficacy and Toxicity outcomes if the underlying, possibly disease-specific, assumptions that are required can be satisfied. Table 2 lists the therapeutic outcome categories, the corresponding true probabilities, and the probabilities estimated from the AA findings. The potentially 12 categories corresponding to the possible combinations of efficacy and toxicity outcomes are collapsed into 6 because materially less efficacy, intolerable toxicity, or worse toxicity without materially better efficacy all identify unacceptable clinical outcomes that do not need to be considered separately.

**Table 2.**
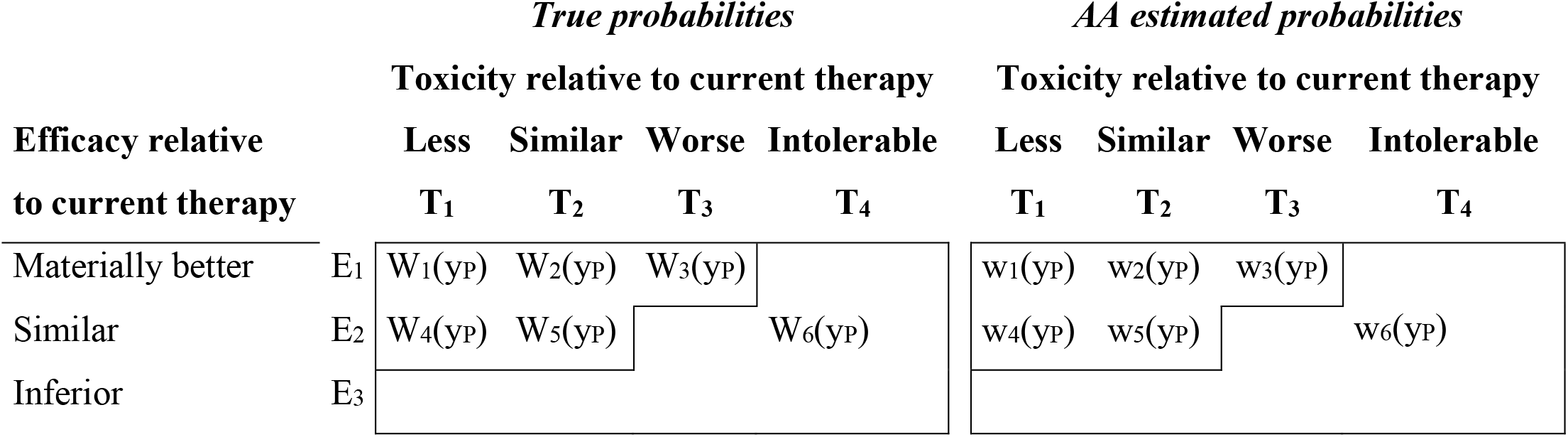
True and estimated probabilities associated with the therapeutic outcomes in Figure 2 and Table 1 for a patient with attributes y_P_.

The estimated probability of the last category [w_6_(y_P_)] may be low because any approval, let alone AA, seems unlikely in that case. However, reality may be different if the intolerable toxicity is a rare event unobserved at the time of AA. The probabilities associated with the combined outcomes generally will depend on both the efficacy and toxicity outcomes. Efficacy and toxicity may reflect a common mechanism of action, and both are often correlated with pharmacokinetic exposure.

### Statistical considerations

Details of how the statistical issues should be addressed are outside the scope of this paper because they will be situation-specific. Nonetheless, sources of uncertainty should be kept in mind when using short-term AA findings based on biomarkers and surrogate measures to predict clinical outcomes. Factors affecting the clinical outcome relevance and predictability for patients include

1. Early trial sampling variance, e.g., the standard error of the early endpoints from AA trials.
2. Selection bias that occurs because AA ordinarily would be granted only when early findings are strong. Some drugs of modest efficacy could by chance provide an early finding strong enough to justify AA, while some drugs of greater efficacy could not provide sufficiently strong early findings, also by chance, and thereby not be granted AA.
3. The correlation between the observations that justify AA and actual clinical outcomes – that is, how well the early findings predict clinical outcome.
4. How well the population represented in the AA trials represents the general population for whom the drug may be prescribed and the circumstances of its clinical use.
5. The consistency of a patient’s predicted clinical outcome from treatment that reflects the variability of the attributes used to predict the outcome and the randomness of the patient’s outcome given a set of attribute values.
6. The variability and sensitivity of the measures of utility to the prior specifications of uncertainty and the effect of the amount of information used for their computation.
7. Decision makers have unequal access to information. Sponsors and FDA have access to details of individual patients in the drug development programs, while other stakeholders (patients, prescribers, HTAs, and payers) do not have this information.
8. Small sample sizes: AA trials tend to include fewer patients than conventional regulatory trials. Consequently, their findings are subject to more uncertainty and may present an overly optimistic picture of treatment effect.

For example, as illustrated in Figure 3, an analysis of 24 month survival in 41 trials of treatments for recurrent glioblastoma found that the smallest trials had the highest survival rates while the larger trials tended to have lower survival rates. [30]

**Figure 3.**
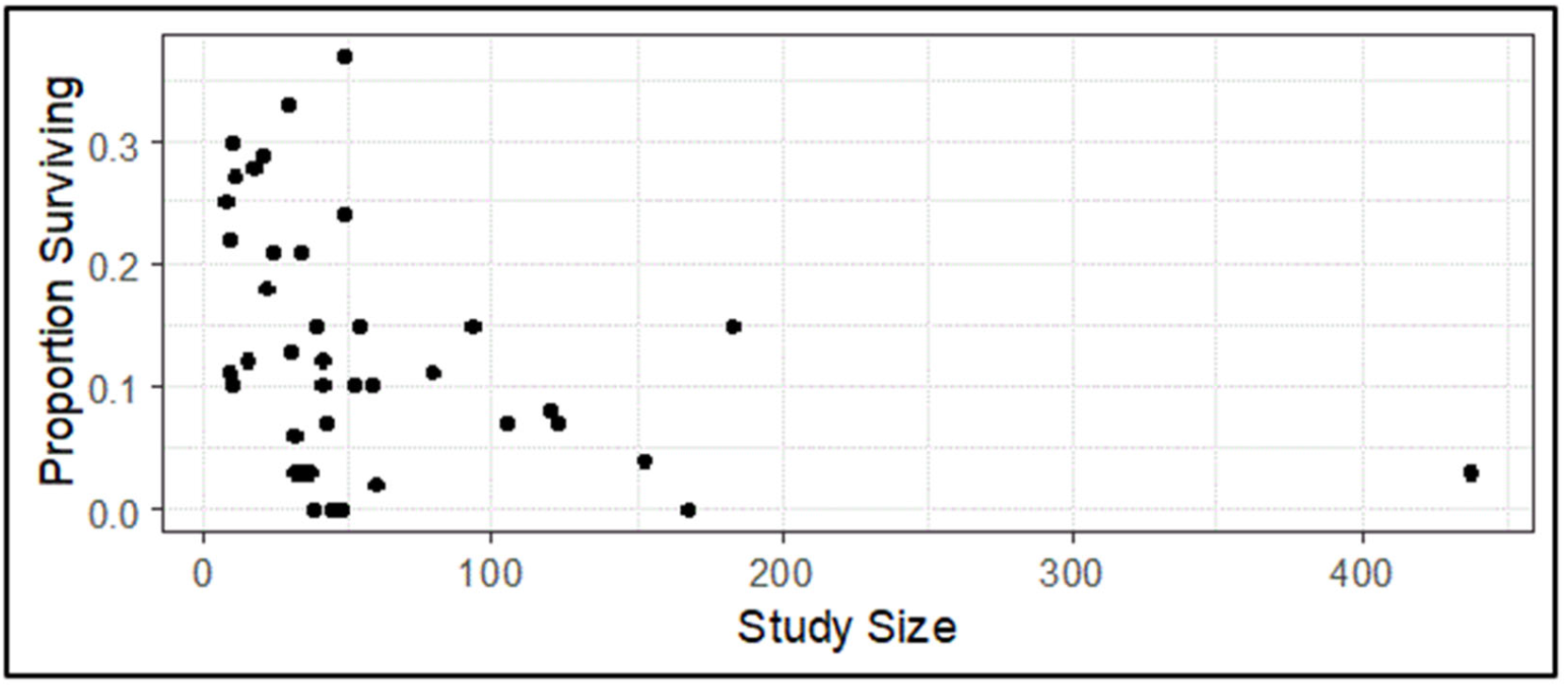
Proportion of patients surviving 24 months or longer in 41 trials of treatments for recurrent glioblastoma.[30].

### Determining the values of the consequences of stakeholder actions

The discussion to this point, summarized in Table 2, describes a way to conceptualize the probabilities associated with the clinical outcomes based on the information provided in the AA trials. However, these probabilities do not by themselves provide sufficient guidance for determining the impact of these trials. This impact depends on how information from the trials drives decisions about possible treatment actions. These decisions also depend on the values the stakeholder attaches to the therapeutic outcome possibilities in Table 2. A numerical measure that reflects these probabilities and values provides a quantitative way to assess impact. This approach is outlined below, and a hypothetical example is presented in Section 3.

At its most basic level, the decision process amounts to generating an action about treatment, e.g.,

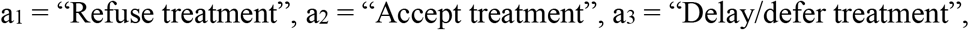

The third possibility, “Delay/defer treatment” allows for some hedging, e.g., to obtain further information about the anticipated effect of treatment by consultation with other specialists, or to consider further the implications about the uncertainty of the predicted outcome due to the limited information from the AA trials. The action taken depends on the information that efficacy and toxicity findings from the AA trials provide and the anticipated relevance for the individual patient. For the present, suppose that the action taken depends on the estimated probabilities for the patient of the various clinical therapeutic outcomes, which may be all that the physician and patient know from the AA findings. [31, 32]

Suppose that a stakeholder can assign a value (‘utility’ in decision analysis terms, or desirability) to the consequence (therapeutic outcome) of any action that the stakeholder might take. The term “value” is used in what follows to indicate a measure of desirability of an action/outcome combination from the standpoint of the stakeholder. The possible therapeutic outcomes reflecting efficacy and toxicity from Table 2 can be considered in the context of the actions from the preceding paragraph (“accept treatment”, “defer/delay treatment”, and “refuse treatment”). The values corresponding to the action/outcome combinations can be expressed as in Table 3, where entry v_ij_ refers to the value that the stakeholder attaches to the therapeutic outcome x_j_ when action a_i_ is taken. The actual v_ij_ values depend on who is specifying the values (e.g., physician, patient) and on the patient’s status. Different stakeholders will have different value tables.

**Table 3.** Typical value table.

| **Action** | **Therapeutic Outcome** |  |  |  |
| --- | --- | --- | --- | --- |
|  | **x<sub>1</sub>** | **x<sub>2</sub>** | **...** | **x<sub>6</sub>** |
| a<sub>1</sub> | v<sub>11</sub> | v<sub>12</sub> | ... | v<sub>16</sub> |
| a<sub>2</sub> | v<sub>21</sub> | v<sub>22</sub> | ... | v<sub>26</sub> |
| a<sub>3</sub> | v<sub>31</sub> | v<sub>32</sub> | ... | v<sub>36</sub> |

Given estimates of the probabilities of the therapeutic outcomes that might be realized as a consequence of taking an action, the value (utility, desirability) of any action can be calculated as the expectation of the values over the set of probabilities corresponding to the therapeutic outcomes.

The possible decisions/actions and outcomes can be defined and agreed to by all stakeholders, though they may not regard these as equally important. Similarly, all of the probabilities can be defined, though stakeholders may not agree on how close these are to the true probabilities (left panel in Table 2). The values, which drive the decisions are likely to be more stakeholder-dependent, and is the focus of the following paragraphs.

The expected value of each action reflects the action-dependent values that a stakeholder (e.g., patient) assigns to each therapeutic outcome, and the stakeholder’s assessment of the likelihood of the outcomes corresponding to the therapeutic options.

Given true and estimated probabilities of the outcomes in Table 2 and the values in Table 3, the <u>true values</u> corresponding to the various actions are

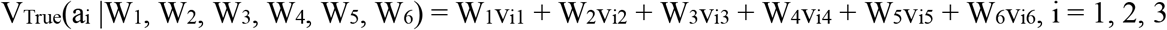

and the <u>estimated values</u> corresponding to the various actions are given by

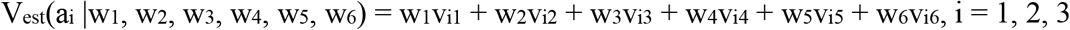

In practice, a patient might choose the action that gave the largest estimated expected value.

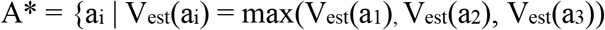

The best choice arguably would be the action a_i_ that maximized the expected value based on the unknown true probabilities of the various outcomes,

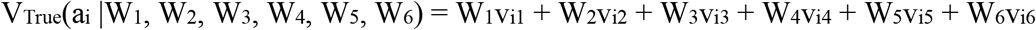

[which could be quite different from V_est_(a_i_ |w_1_, w_2_, w_3_, w_4_, w_5_, w_6_)], so that

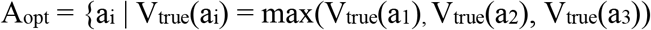

The optimal action A_opt_ depends on the true probabilities. The value of the “best” course of action that the patient or physician conceivably could take based on the AA finding is V(A_opt_), the metric value associated with A_opt_. Unfortunately, this value is not known because the true clinical outcome probabilities on which it is based (W_1_ – W_6_) are unknown.

We focus on actions that the evidence in hand suggests provides the best value. What constitutes a “best” value for a stakeholder depends on the stakeholder’s value table. Although, for example, a patient and a physician should have the same outcome probabilities based on the findings from the AA trial(s) and the true “State of Nature”, they may have different value tables (Table 3) and so may assess the impact of the AA trials differently. Consequently, their best choice actions may differ, and will need to be reconciled so that a decision about the therapeutic course can be made. Framing the process as just described should facilitate reaching a decision because the focus is concentrated on the values, which can be the basis for discussion.

All stakeholders do similar calculations, but with different value definitions because of differing perspectives. This process described here focuses on the integration of the value matrix differences, which could be subjective, and not on the conclusions from the AA findings, which are not subjective (or at least are less subjective). This issue has been considered in the literature.[33, 34]

The different value arrays of the various stakeholders represent the values they assign to the possible actions they might take. As noted in the introduction, these potential actions will differ among stakeholders. This circumstance arises in the design of confirmatory trials based on biomarkers and when different subgroups of the general population might have different clinical outcomes from a particular therapy. Decision theoretic principles can be useful in addressing the issues that arise in these contexts. [14, 17, 18]

The willingness of different stakeholders to accept uncertainty might not be aligned even though seemingly the same evidence is assessed. Reaching consensus on the value of a new course of treatment is a non-trivial task that requires careful attention to the underlying issues. [6, 8] A process that requires stakeholders to explicitly state probabilities and expectations may facilitate better recognition and understanding of these conflicts. Having a common process (and metric) should help discussions of policy and strategy with respect to AA.

For example, HTAs and payers often require more complex studies that allow a robust prediction of the future likely use of a new drug in the context of diverse national healthcare systems. AA development programs should consider the possibility that the shortcomings of the AA process could hamper HTA assessments. This means that discussion and dialogue with HTA/Payer organizations as well as regulators aimed at prior consensus or agreement should be undertaken before embarking on an AA development program.

## Illustrative Example

The following simple example illustrates the basic ideas of the proposed method. Its application in practice could be appreciably more complex.

### Findings from an AA trial

Suppose that Accelerated Approval is based on the findings from a single-arm trial of 50 or perhaps 100 patients carried out in a homogeneous patient population to evaluate the effect of combining a new treatment with “standard of care” (SoC) in the treatment of a rare cancer for which there is no effective treatment. The AA findings may be based on whether a surrogate endpoint such as a biomarker demonstrates positive finding known not to occur with SoC. Let P denote the true, unknown probability that a patient receiving the new treatment demonstrates a positive surrogate endpoint effect (SE). The trial will be considered a success if the outcome of the trial demonstrates that P is likely to exceed 20%. Toxicity also may be an issue, so an estimate of the value of Q, the probability that a patient receiving the new treatment experiences a severe toxic event, also will be obtained.

The analysis of the AA trial could be carried out using a conventional frequentist or a Bayesian approach. The Bayesian approach will be used for the purpose of illustration; conventional frequentist calculations yield similar results. Suppose, *a priori*, that one expects P to be about 10% and Q to be about 20%. These are pessimistic assumptions because the success of the trial depends on demonstrating that P is larger than 0.20. For the purpose of analysis, the prior distributions of P and Q are taken to be beta distributions with parameters (1, 9) for P and (2, 8) for Q; the expected values of these distributions are 0.10 and 0.20, respectively. Since the trial is relatively small, the effect of sampling variability of the estimates of P and Q based on the outcome of the trial needs to be taken into account, e.g., by credible (or confidence) intervals.

Suppose first that N = 50, 15 of the 50 patients had a positive SE effect, and none had a severe toxicity event. This means that the posterior distribution of P is a beta distribution with parameters (1+15, 9+35) = (16, 44). Consequently, the posterior expected value of P is 16/60 = 0.27, a 95% credible interval for P is (0.16, 0.38), and the posterior probability that P > 20% is 0.88. This could be regarded as a successful outcome if being 88% ‘sure’ that P > 20% defines ‘success’. The posterior distribution of Q is a beta distribution with parameters (2, 58) so that the posterior expected value of Q is 2/60 = 0.03 and a 95% credible interval for Q is (0.004, 0.09).

If N = 100 with 30 of the patients having a positive SE effect and none having a severe toxicity event, then the posterior distribution of P is a beta distribution with parameters (31, 79) so that the expected value of P is 31/110 = 0.28, a 95% credible interval for P is (0.20, 0.37), and the posterior probability that P > 20% is 0.98, strongly suggesting that the trial had a ‘successful’ outcome. The posterior distribution of Q is a beta distribution with parameters (2, 108) so that the expected value of Q is 2/110 = 0.018 and a 95% credible interval for Q is (0.002, 0.05). That is, given the trial outcome, the expected risk of toxicity is about 2%, not 20% as assumed at the outset.

### Predicting clinical outcome

Suppose that this finding is sufficiently promising, and the need for therapy sufficiently dire, so that AA may be granted for adding the new treatment to SoC. A key issue at this point is the reliability of the clinical outcome predictions based on the trial findings, as outlined in Table 2. The SE may or may not turn out to be a good surrogate for a tangible clinical benefit (CB) such as surviving for 6 months or for 5 years for patients with this cancer. Different stakeholders may attach quite different values to the importance of the SE as a predictor of the probability of demonstrating CB.

In reality, demonstrating a positive SE does not guarantee CB nor does failure to demonstrate a positive SE rule out the possibility of CB. Suppose for the sake of illustration that a patient demonstrating a positive SE has a 70% chance of demonstrating CB with treatment, i.e., Prob(CB|SE) = 0.7. Suppose also that a patient who does not demonstrate a positive SE has a 100q% chance of achieving CB with treatment, i.e., Prob(CB|no SE) = q. CB, being the best outcome, corresponds to efficacy outcome E_1_ in Figure 2. If P(SE) = 0.28, then the probability that a treated patient achieves CB is

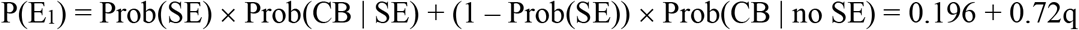

We assume for illustration that achieving CB without demonstrating SE is arguably unlikely if the surrogate effect is based on a biomarker that has been demonstrated to have experimental validity. This means that for this example, the value of q is likely to be small, say less than 0.15, so that a patient who does not demonstrate a positive SE effect with treatment has somewhere between a 0% and a 15% chance of demonstrating CB. With this assumption, the estimated probability of achieving CB, 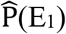, lies between 0.20 and 0.30. For illustration, assume q = 0.05, so that 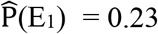 when Prob(SE) = 0.28.

Inferiority of clinical effect is irrelevant in this scenario because there are no effective treatments. Consequently, 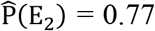 and 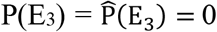. With respect to toxicity, adding a treatment with a potentially toxic effect cannot decrease the probability of severe toxicity, so T_1_ cannot occur i.e., P(T_1_) = 0. The expected toxicity rate based on the AA trial is 3%, so 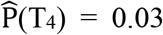. Consequently, 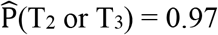. For illustration, suppose that 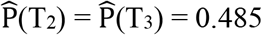.

Again, even though it is not necessarily realistic, nor is it assumed for Table 2, suppose for simplicity of exposition that efficacy and toxicity outcomes are independent. Then the probabilities in Table 2 can be summarized in Table 4 for the various cases. The expected values of P and Q are as described above. The Worst Case values (pessimistic assessment) consist of the lower bound for P and the upper bound for Q, while the Best Case values (optimistic assessment) consist of the upper bound for P and the lower bound for Q.

**Table 4.** Predictive probabilities of therapeutic outcomes as a function of anticipated values of P (probability of positive SE) and Q (probability of severe toxicity) when 15 out of 50 patients in a trial demonstrate SE and no patients demonstrate severe toxicity. The values corresponding to a sample of 100 patients with 30 demonstrating SE are nearly the same. P(E_1_) = estimated probability of clinical benefit (CB)

|  |  |  |  | Predicted Therapeutic Outcome |  |  |  |  |  |  |
| --- | --- | --- | --- | --- | --- | --- | --- | --- | --- | --- |
|  |  |  |  | Efficacy | Better | Better | Better | Similar | Similar |  |
|  |  |  |  | Toxicity | Less | Similar | Manageable | Less | Similar | Other |
|  | P | Q | p(E1) |  |  |  |  |  |  |  |
| Expected | 0.27 | 0.03 | 0.23 |  | 0 | 0.11 | 0.11 | 0 | 0.38 | 0.41 |
| Worst Case | 0.16 | 0.09 | 0.15 |  | 0 | 0.07 | 0.07 | 0 | 0.41 | 0.44 |
| Best Case | 0.38 | 0.004 | 0.30 |  | 0 | 0.14 | 0.14 | 0 | 0.34 | 0.37 |

The entries in Table 4 correspond to the estimated probabilities in Table 2 and assumptions about the predictability of CB from SE, and illustrate the effects of sampling variability and AA trial sample size. The true probabilities of the various therapeutic outcomes are needed to assess the impact of AA. Table 5 provides the true predicted probabilities of the therapeutic outcomes for each of six illustrative scenarios exploring the effect of various values of the probabilities of SE and a toxic effect.

**Table 5.**
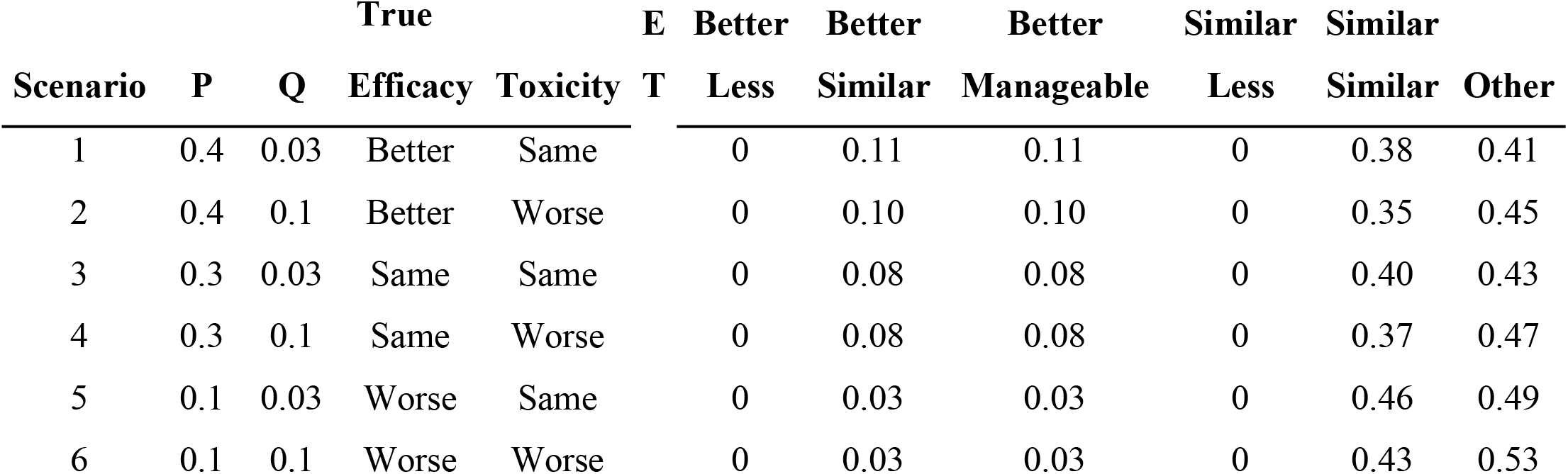
Illustrative scenarios for values of P = Prob(SE) and Q = Prob(Toxicity)

### Values corresponding to possible actions

In this example, “similar efficacy” is the same as “no efficacy” because there are no effective treatments. Likewise, “refusing treatment” is the same as ‘deferring/delaying treatment”. Table 6 provides two sets of values for the entries in Table 4. These two value sets could correspond to the values for two different patients or to a patient and the physician guiding the patient’s treatment.

**Table 6.**
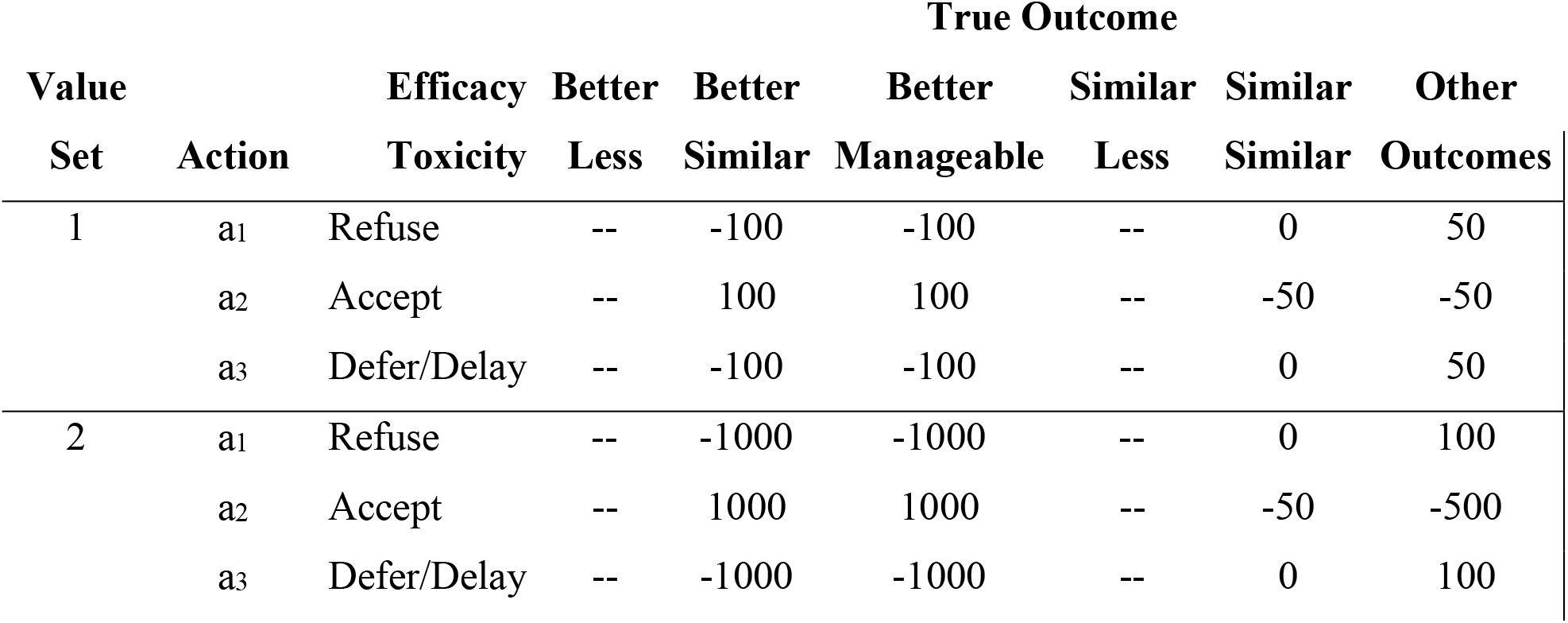
Value table for example.

Accepting treatment when the efficacy is not “better” has a negative value because there is a potential economic issue due to the cost of the new medication. If the “True Outcome” (State of Nature) is “Better Efficacy and Less Toxicity”, then the patient (and the physician) might set the value of accepting treatment (a_2_) at 100 or maybe 1000 because it’s good to get the better treatment. There is, of course, no necessity for the patient and the physician to have the same value tables.

If, on the other hand, the “True Outcome” is anything other than at least similar efficacy and no worse than manageable toxicity, then the patient might set the value of accepting treatment at -50, because getting a worse or very intolerable treatment would be unacceptable. The value of refusing treatment (a_1_) if the treatment offers better efficacy and less toxicity might be -100 because it might be tragic to miss an important therapeutic opportunity. Finally, refusing or deferring treatment might imply that the value should be 100 or 1000 because it would be highly desirable to avoid intolerable toxicity or opportunity loss from ineffective treatments.

Table 7 combines the values (2 sets) in Table 6 with the estimated and true therapeutic outcome probabilities in Tables 4 and 5 to give the estimated and true values corresponding to the various scenarios, choices of bounds, and sample sizes. This table provides the basis for assessing how a patient (or physician) might prefer a course of action.

**Table 7.** Expected utilities (values) corresponding to alternative courses of action as a function of the outcome of the AA trial and the unknown true probabilities of achieving a positive SE effect or experiencing severe toxicity when N = 50. The values for N = 100 are similar.

| (P, Q values) | Value Set 1 |  |  | Value Set 2 |  |  |
| --- | --- | --- | --- | --- | --- | --- |
|  | Action (Decision) |  |  | Action (Decision) |  |  |
|  |  |  | Defer/ |  |  | Defer/ |
|  | Refuse | Accept | Delay | Refuse | Accept | Delay |
| Expected | 7.5 | -28.1 | 7.5 | -101.9 | -96.0 | -101.9 |
| Worst Case | 17.1 | -37.8 | 17.1 | -31.1 | -191.3 | -31.1 |
| Best Case | -1.4 | -18.2 | -1.4 | -172.3 | -5.7 | -172.3 |
| True Scenario 1 | -1.4 | -17.4 | -1.4 | -176.6 | -4.7 | -176.6 |
| True Scenario 2 | 2.3 | -19.8 | 2.3 | -156.7 | -40.5 | -156.7 |
| True Scenario 3 | 5.4 | -25.6 | 5.4 | -119.6 | -74.0 | -119.6 |
| True Scenario 4 | 8.6 | -27.3 | 8.6 | -103.8 | -104.7 | -103.8 |
| True Scenario 5 | 19.0 | -41.9 | 19.0 | -5.5 | -212.5 | -5.5 |
| True Scenario 6 | 21.2 | -42.4 | 21.2 | 2.1 | -233.2 | 2.1 |

The values of P and Q in any of the cases or under any of the True Scenarios are not under the control of the patient or physician (or, in general, of any stakeholder). The stakeholders control the values associated with each decision/therapeutic outcome combination, and these values are crucial for identifying appropriate decisions. A striking finding from the Value Set 1 part of Table 7 is that accepting treatment would not be the best course of action essentially regardless of the values of P and Q assumed here, so that the treatment is unlikely to be useful or acceptable to the patient. This could mean, in evaluating the treatment for AA, that AA might not be appropriate at all. However, increasing the value of a CB (Value Set 2) implies that accepting treatment could lead to the “largest” utilities and therefore be a preferred decision under the “Expected” and “Best Case” values of P and Q or True Scenarios 1-3 even if a CB is unlikely.

That is not to say, of course, that if Value Sets 1 and 2 corresponded to the values of a patient and a physician, reflecting what each thought was important, that the action that the physician would recommend would be the same as the action that the patient’s analysis revealed (or the same as what the analysis of any other stakeholder, e.g., an HTA) revealed. Resolving the differences would require discussion and negotiation, but ideally could be simplified because all of the stakeholders were using the same metric.

Of course, decision makers do not know which scenario actually applies in any real situation. One way to resolve the uncertainty as to which scenario to accept and, therefore, which treatment option to take, would be to use historical information from the AA trial(s) or other sources if available to estimate probabilities corresponding to the true scenarios. The confidence one might have in the reliability of the decision certainly would depend on the strength of the historical evidence, including the size of the AA trial(s), as indicated in section 2.4.

## Discussion

In a 21^st^ century clinical development paradigm, the AA process represents an avenue for bringing potentially life-saving new medical treatments to patients sooner. However, drugs approved via the AA approach may turn out not to provide actual clinical benefit. AA s generally are based on fewer, smaller, or shorter clinical trials than are used to support conventional regulatory approval, so that there will be less information about the occurrence of rare or delayed adverse events that may emerge over time and affect the label.[35] The uncertainty about the actual clinical value of a new treatment receiving regulatory approval based on surrogate endpoints is the risk that must be accepted in order to have expedited availability of new products that patients can use as prescribed by their medical providers. Product performance may become increasingly more difficult to evaluate as a result of a provision in section 3022 of the 21^st^ Century Cures Act that requires the FDA to consider lower standards of evidence, including ‘real world evidence”.

Under AA, health system stakeholders need to be able to communicate with each other using standardized metrics so that the health value of the new treatment that has just been approved can continue to be evaluated for its meaningfulness as the treatment sponsor continues clinical development activities. The principles described above are an attempt to formulate a process for the evaluation. Such a process is needed and important because all stakeholders (patients, physicians, sponsors of the medical treatment being developed, regulatory authorities and payers) need to understand how the new product performs for patients for whom the product is indicated.

The framework proposed here differs from decision science based methods that have been described previously in that stakeholders decide on a course of action that reflects stakeholder-specific values by an objective, transparent, and reproducible process that differs among stakeholders only with respect to the stakeholder-specific actions and values. Stakeholders could reach different action decisions, possibly at different points in time, so that there may be no immediate determination of ‘consensus’ (as opposed to the decision model described by Xu *et al* [13]). It also has a different aim from methods that focus on sponsor decisions about confirmatory or post-POC study designs, and how these design options could affect outcomes, utility, and future (post-approval) decisions by stakeholders. The context differs as well from previous work in that not all stakeholders implement their decision processes contemporaneously because the AA process pushes important decisions from the sponsor and regulatory agencies to stakeholders that determine implementation in clinical care.

Although the proposed process is general, its implementation provides a starting point on which the health systems stakeholders can focus while preparing their own internal systems for capturing the relevant information necessary in a paradigm where the product is now available to the public based on AA. The current regulatory paradigm has been evolving to address criticisms that it has been holding up the ability of potentially lifesaving treatments to reach patients for whom effective treatment alternatives do not exist. The ideas presented here describe a process for facilitating the effectiveness of the paradigm as it evolves by providing value added communication among all stakeholders.

This description of the process for assessing the impact of AA has omitted many implementation details in the interest of clarity and brevity that we anticipate addressing in future articles. For example, the interpretation of the outcomes of AA trials and their use to attach probabilities to predicted outcomes depends on the attributes and design of the AA trials such as whether the trials had 1 or 2 arms, the nature of the controls, if any (e.g., active, standard of care, placebo/no control), the nature of the response (dichotomous, discrete, continuous), etc. Additional details include the models used to obtain predictive distributions for clinical outcomes based on the outcomes of AA trials and historical information, models for regulator-sponsor interdependence, continuous outcomes, and the effect of dependence of efficacy and toxicity outcomes. Moreover, the biology of the underlying disease may be influential in ascertaining the true impact of AA.[36, 37]

Greater uncertainty about perceived treatment effectiveness may increase the hesitancy of payers to provide reimbursement to patients choosing to receive new treatments. Sponsors may accept greater uncertainty than would be optimal from a public health perspective.[17, 18] Payers will often use the concept of number needed to treat (NNT) or number needed to harm (NNH) to evaluate this uncertainty to make reimbursement decisions.[38, 39] Other considerations also are important, such as the potential Incremental Cost Effectiveness Ratio of a new treatment compared with a “best” alternative treatment based on key efficacy variables and the cost of approved screening tests for targeted therapies.

## Data Availability

All data produced in the present work are contained in the manuscript

## Acknowledgements

This work is a collaborative effort of members of the DIA Innovative Design Scientific Working Group and the Expedited Approvals subgroup of the DIA Bayesian Statistics Working Group. We express our appreciation to the members of the working groups for their insights and contribution to the discussions of the many revisions of the original concept that are expressed in this article: Matt Austin, Cong Chen, Jennifer Clark, Vladimir Dragalin, Zhaowei Hua, Qi Jiang, Sandeep M. Menon, David Norris, Nitin Patel, Martin Posch, Jane Qian, Kiichiro Toyoizumi, Ling Wang, and David Wright.

## Appendix 1 Accelerated Approval

Accelerated Approval is one of four programs identified in the current FDA guideline aimed at facilitating and expediting the development and review of new drugs to address unmet medical needs for treating serious or life threatening conditions.[1] The following table presents an abbreviated summary of the four programs.

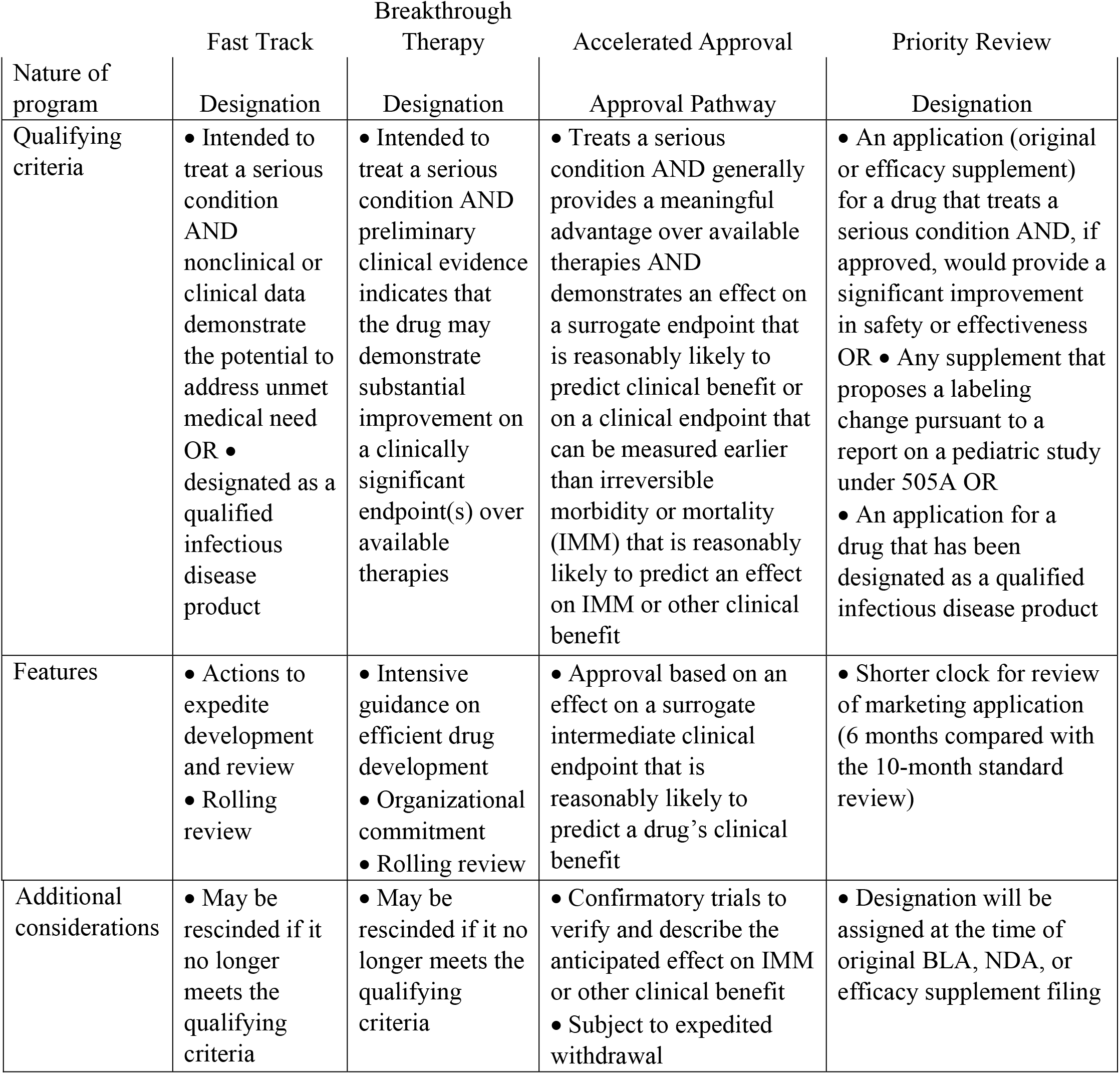

| Nature of program | Fast Track | Breakthrough Therapy | Accelerated Approval | Priority Review |
| --- | --- | --- | --- | --- |
|  | Designation | Designation | Approval Pathway | Designation |
| Qualifying criteria | Intended to treat a serious condition AND nonclinical or clinical data demonstrate the potential to address unmet medical need<br>OR • designated as a qualified infectious disease product | Intended to treat a serious condition AND preliminary clinical evidence indicates that the drug may demonstrate substantial improvement on a clinically significant endpoint(s) over available therapies | Treats a serious condition AND generally provides a meaningful advantage over available therapies AND demonstrates an effect on a surrogate endpoint that is reasonably likely to predict clinical benefit or on a clinical endpoint that can be measured earlier than irreversible morbidity or mortality (IMM) that is reasonably likely to predict an effect on IMM or other clinical benefit | An application (original or efficacy supplement) for a drug that treats a serious condition AND, if approved, would provide a significant improvement in safety or effectiveness<br>OR • Any supplement that proposes a labeling change pursuant to a report on a pediatric study under 505A<br>OR • An application for a drug that has been designated as a qualified infectious disease product |
| Features | Actions to expedite development and review<br>Rolling review | Intensive guidance on efficient drug development<br>Organizational commitment<br>Rolling review | Approval based on an effect on a surrogate intermediate clinical endpoint that is reasonably likely to predict a drug's clinical benefit | Shorter clock for review of marketing application (6 months compared with the 10-month standard review) |
| Additional considerations | May be rescinded if it no longer meets the qualifying criteria | May be rescinded if it no longer meets the qualifying criteria | Confirmatory trials to verify and describe the anticipated effect on IMM or other clinical benefit<br>Subject to expedited withdrawal | Designation will be assigned at the time of original BLA, NDA, or efficacy supplement filing |

The Accelerated Approval program applies to drugs that treat serious conditions and fulfill an <u>unmet medical</u> need based on a <u>surrogate endpoint</u>. A <u>surrogate endpoint</u> is a marker, such as a laboratory measurement, radiographic image, physical sign or other measure that is thought to predict clinical benefit, but is not itself a measure of clinical benefit. An application for accelerated approval should also include evidence that a proposed surrogate endpoint or an intermediate clinical endpoint is reasonably likely to predict the intended clinical benefit of a drug. The evidence that provides support for the use of a surrogate endpoint will depend in the circumstances but usually will be based on considerations such as whether they measure the underlying cause of the disease, an effect that predicts the ultimate outcome, or the state of the pathophysiologic pathway leading to the clinical outcome. Epidemiologic evidence can be useful here, but requires an assessment of whether there is reliable and consistent epidemiologic evidence supporting the relationship between the endpoint and the intended clinical benefit, how precisely the epidemiologic relationship between the endpoint and clinical outcome is defined, and whether the effect on the surrogate endpoint has been shown to predict a clinical benefit with another drug or drugs.

A <u>serious condition</u> is a disease or condition associated with morbidity that has substantial impact on day-to-day functioning that clinical judgment suggests will progress in severity or lead to mortality if left untreated. The drug in question must be intended to have an effect directly or indirectly on a serious condition, including mitigating or preventing a serious treatment-related side effect or avoiding or diminishing a serious AE associated with available therapy.

An <u>unnmet medical need</u> is a condition whose treatment or diagnosis is not addressed adequately by available therapy. If there is no available therapy for a serious condition, there is clearly an unmet medical need. A new treatment could be considered to address an unmet medical need even when available therapy exists, under certain conditions. [1]

The guidance states that FDA may grant accelerated approval to a product for a serious or life-threatening disease or condition upon a determination that the product has an effect on a surrogate endpoint that is <u>reasonably likely</u> to predict clinical benefit, or on an intermediate clinical outcome ascertainable sooner than irreversible morbidity or mortality that is <u>reasonably likely</u> to predict an effect on irreversible morbidity or mortality. The accelerated approval pathway has been used primarily in settings in which the disease course is long and an extended period of time would be required to measure the intended clinical benefit of a drug. Accelerated approval also may be useful in acute disease settings where the clinical event for which benefit would be realized occurs rarely so that very large trials would be needed to demonstrate benefit.

The 2016 21^st^ Century Cures Act that mandated the establishment of programs for expedited approval of these drugs also required that that the FDA develop ‘patient-focused drug development guidance’ that addresses how the FDA plans to use patient experience data ‘with respect to the structured risk-benefit assessment framework’ described in the Federal Food, Drug, and Cosmetic Act, while adhering to the (substantial) evidence standard required by the FFDC Act. [40] The principal risk of the accelerated approval approach is the possibility that patients will be exposed to a drug that ultimately will not be shown to provide an actual clinical benefit. Also, there generally will be fewer, smaller, or shorter clinical trials than is typical for a drug receiving traditional approval, which usually will mean there is less information about the occurrence of rare or delayed adverse events that subsequently will emerge and be incorporated into the label.[35]

Consequently, d<u>rug companies still are required to conduct studies to confirm the anticipated clinical benefit</u> with due diligence and promptly. In general, the confirmatory trial would evaluate a clinical endpoint that directly measures clinical benefit, ordinarily in the same disease population that was studied to support accelerated approval. If these confirmatory trials show that the drug actually provides a clinical benefit, then the FDA grants traditional approval for the drug. If they do not show that the drug provides clinical benefit, FDA has (in principle) regulatory procedures in place that could lead to removing the drug from the market.

The Accelerated Approval process has been in place for a number of years. A recent study evaluated the preapproval and confirmatory trials of drugs granted accelerated approval between 2009 and 2013. [41] Accelerated approval was granted to 22 drugs for 24 indications (19 for treating cancer), based on a total of 30 preapproval studies. Eight of the studies included fewer than 100 participants, and 2/3 (20) of the studies included fewer than 200 participants. After 3 or more years of followup, half (19 of 38) of the required confirmatory studies were completed. Most importantly, clinical benefit had not been confirmed for 8 indications approved at least 5 years previously.

## Appendix 2 Elements of a Statistical Decision Framework

1. <u>Data</u> consisting of a variable X taking values in **𝒳** = {x_1_, x_2_, …} or some interval on the real line, etc. with a likelihood f_X_(x; θ) depending on parameter(s) θ ≡ “State of Nature”.
2. A <u>prior distribution</u> g(θ;Ψ), θ ∈ Θ, for the unknown true value of θ; Ψ = parameter(s) of the prior distribution of θ.
3. A <u>decision procedure</u> D(x) that identifies an <u>action</u> to be taken on observing X = x; D(x) is a member of a set **𝒟** = {D_1_, D_2_, …} of possible decision procedures. This is key because the point of obtaining the data is to provide guidance for choosing an appropriate action.
4. A set of <u>possible actions</u> A = {a_1_, a_2_, … } consisting of the possible values of D(x). Different decision rules could specify different actions even with the same data.
5. A <u>rule</u> V(D(x), θ) that assigns a <u>value</u> to the action (in A) specified by D(x) when the true value of the State of Nature is θ. This rule has various names, e.g., loss function, utility function, etc. Assume for convenience that the rule is a loss function, so that small values are desirable. This relates a value to an action that the decision function D(x) specifies when x is given.
6. A <u>risk function</u> R(D,θ) corresponding to D that is the expected value of V with respect to x given θ,

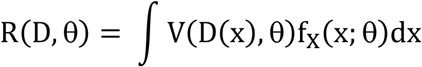

(this would be a summation if the values in X are discrete). Every decision procedure D(x) has its own range of risks corresponding to the various “States of Nature” (values of θ). The risk value for any decision procedure D(x) reaches a maximum for some θ ∈ Θ. This depends on the random process that generates x values (i.e., the value of θ), but not on any particular value of x.
7. A <u>minimax</u> (or <u>admissible</u>) decision procedure D* is the decision procedure in **𝒟** that has the smallest value of the maximum risk over the set of values of θ ∈ Θ; more formally,

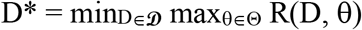
8. The <u>Bayes risk</u> r_g_(D; Ψ) of a decision procedure D is the expected value of the risk function R(D, θ) with respect to the prior distribution of θ,

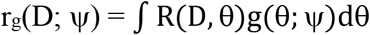
9. A <u>Bayes Rule</u> is the decision rule D^#^ in **𝒟** that that provides the smallest value of r_g_(D; Ψ) so that

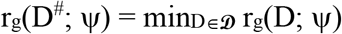

Every admissible procedure is a Bayes Rule.

